# Intelligence as proxy phenotype providing insight into the heterogeneity of schizophrenia

**DOI:** 10.1101/2023.02.04.23285470

**Authors:** Hongyan Ren, Meiyu Yin, Qiang Wang, Wei Deng, Xiaohong Ma, Liansheng Zhao, Xiaojing Li, Pak Sham, Ming Li, Shiwu Li, Tao Li

## Abstract

Schizophrenia is a mental disorder constituting different symptom clusters. Its high heterogeneity in both pathophysiology and clinical manifestations hampered effective prevention and treatment. It has long been recognized that one of the core features of schizophrenia is its intellectual decline. Using the proxy-phenotype method (PPM), we tried to identify core genes, the expression of which in the dorsal lateral prefrontal cortex (DLPFC) showed a genetic dependence between intelligence (IT) and schizophrenia (SCZ). The result revealed ten genes of genetic dependence in their genetic expression in DLPFC between IT and schizophrenia. Further, a clustering analysis using the expression matrix of these ten genes identified four biotypes in our patient group. Subsequent phenotypic profiling of these four biotypes indicated a significant difference in working memory capacity, the gray matter volume (GMV) of five brain regions (lLimbicA_TempPole_2, rLimbicA_TempPole_2 rLimbicB_OFC_1, rContA_IPS_1 and rContB_PFClv_1), structural network and psychopathology. An in-vitro investigation of the biological functions of these core genes indicated their potentially critical role in neuronal growth, especially in dendritic spines. Our current study employed a novel statistical approach to identify the core genes associated with IT and explore the possibility of using the expression knowledge of these core genes to reduce the heterogeneity of schizophrenia. The results pinpointed one biotype that exhibited significant deficits in working memory, GMV in limbic and prefrontal areas, and also showed psychopathology of core negative symptom and worse outcomes.

## 1. Introduction

Schizophrenia (SCZ) is a complex disease manifesting in the form of syndromic symptoms [1]. Although both twin and familial studies showed a relatively high heritability of about 0.8 [2], it remains elusive to identify all the genetic variants which could explain such a heritability. Reasons for the dilemma include the complex genetic architecture underlying schizophrenia, involving many variants of small to modest effect size (polygenicity) [3] or even all genes in the relevant brain regions (omnigenicity) [4], and the heterogeneity of clinical manifestations.

Since the age of Emil Kraepelin, one of the most recognized theories regarding the nature of schizophrenia is the one of cognitive decline [5]. The cognitive deficits in schizophrenia are pervasive and wide-ranging, from memory [6] and learning [7] to social cognition [8]. Multiple studies adopting different methodologies pointed to the dorsal lateral prefrontal cortex (DLPFC) as the core region in cognitive functioning [9]. The patients with DLPFC lesions scored lower in general intelligence, executive function and working memory than their healthy counterparts [10, 11]. Batiuk et al.carried out a high-resolution characterization of cell types associated with schizophrenia by conducting single-nucleus RNA sequencing on the DLPFC from Brodmann area 9 of patients with SCZ and matched controls. The results showed a general decrease in GABAergic interneurons in schizophrenia, affecting subtypes from all families, particularly those belonging to parvalbumin, SST and VIP, and an increase in some specific principal neurons. A subsequent spatial analysis of altered cell types indicated that the most notable changes took place in the upper-layer subtype of SST-SST CALB1 [12].

Although cognitive deficits subserved by altered neuronal composition and gene expression of DLPFC play a critical role in schizophrenia, the results fall short of clinical translation and generalization due to the high heterogeneity of clinical manifestations of schizophrenia. So far, the studies on schizophrenia still use categorical, phenomenology-based criteria to recruit participants. Nevertheless, many studies have implied that the different clinical syndromes might have different genetic architecture and subsequent pathoetiology [13, 14]. Detangling these clinical syndromes based on their distinctive biological mechanisms could improve the accuracy of diagnosis and facilitate the identification of new treatment agents. However, it is still time-consuming and cumbersome to collect samples based on the specific clinical symptoms of SCZ for well-powered studies. Therefore, Rietveld *et al*. tried to reduce the heterogeneity of SCZ and focus on specific aspects of SCZ by using an alternative approach of the proxy-phenotype method (PPM) [15]. PPM combines the association results of two phenotypes and highlights the loci that show a strong genetic dependence between them. For instance, Bansal *et al*., in their study, combined the GWAS results of education attainment and SCZ and, by using PPM, identified new loci that failed to discover in the original SCZ GWAS [16]. Although their findings are promising, the study used information from genetic variants, which lack spatial and temporal resolution. Therefore, we applied the PPM to the transcriptomic association results of IT and SCZ in DLPFC, aiming to identify core genes showing genetic dependence between IT and SCZ and leverage these core genes’ expression values to delineate new biotypes in schizophrenia.

## 2. Materials and methods

### 2.1. Identification of intelligence (IT)-proxy genes for SCZ

Built upon the study by Bansal et al. [16], we took a proxy-phenotype approach to detect the dependence in gene expression between intelligence (IT) and schizophrenia (SCZ). While genetic correlation is defined by the correlation of the (true) effect sizes of genetic variants on the two traits, genetic dependence means that the genetic variants associated with IT are more likely to also be associated with SCZ than expected by chance [15, 16].

Essentially, the proxy-phenotype method (PPM) is a two-step approach. As demonstrated in Figure 1, firstly, using MetaXcan [17] and summary statistics of IT GWAS, we predicted the transcriptomic profiles in DLPFC for a sample of 363,502 individuals and tested their associations with IT. Secondly, taking 104 genes with a significant association with IT after controlling for the multiple comparisons (0.05/10312 = 4.85 × 10^−6^) to the next step, we identified IT_proxy genes for SCZ by investigating their association with SCZ in a sample of 35,802 individuals consisting11,260 patients and 24,542 controls. Due to the reduced number of tested genes, the significance threshold became less stringent (0.05/104 = 0.00025).

**Fig. 1:**
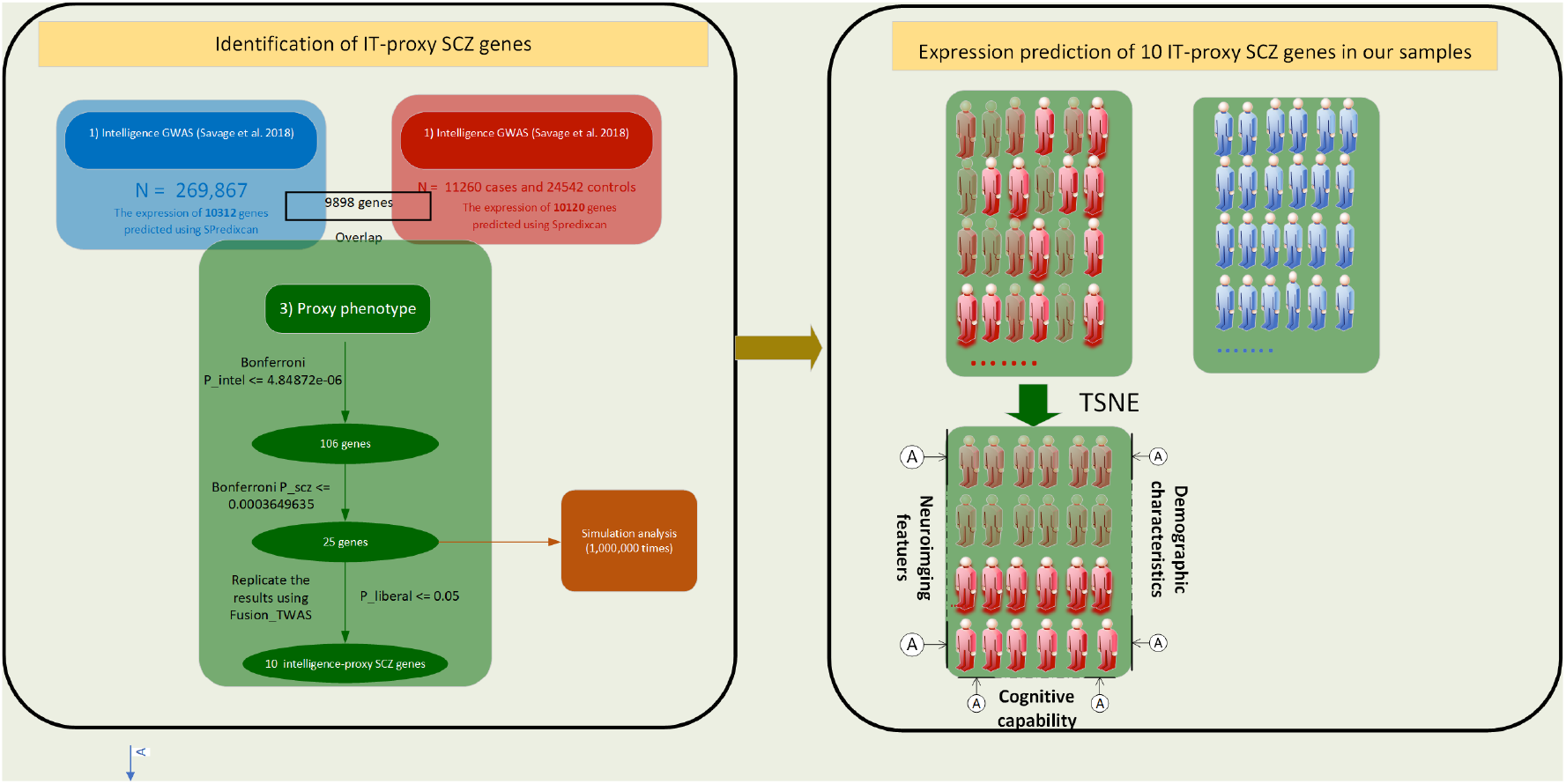
Schematics of analysis pipeline of current study

We then carried out an enrichment analysis for the IT-proxy SCZ genes. To ensure the rigour of identified genes, we simulated the enrichment analysis 10,00,000 times to ensure the significance not expected by chance and chose another program, FUSION TWAS [18], to replicate the results arising from MetaXcan, with a gene being replicated if the association using FUSION TWAS reached a nominal significance.

### 2.2. Expression prediction of IT-proxy SCZ genes

Ten genes arose from the analyses described above. To investigate the phenotypic consequences of these ten IT_proxy genes, including the potential of identifying subtypes in schizophrenia, we used PrediXcan [19] to predict their genetically regulated expression (GReX) in DLPFC in an independent sample including 170 first-episode and drug-naive patients with schizophrenia and 99 healthy controls.

Following the GReX prediction, we subjected the expression matrix (155 individuals, ten genes) to t-Distributed Stochastic Neighbour Embedding (t-SNE) analysis and chose from a range of hyper-parameter (“perplexity”) the one which could separate the patients into the most visually distinctive clusters as the optimal hyper-parameter.

To profile each cluster (biotype), we compared them in demographic features, cognitive performance using CANTAB (https://www.cambridgecognition.com/cantab/), grey matter volumes and psychopathology. Details of cognitive tests and MRI neuroimaging scanning were described in the supplementary materials. The analyses comparing biotypes in demographics, cognitive capabilitiesand gray matter volumes (GMVs) were performed in the R environment; the analysis of longitudinal clinical data was carried out using STATA 17. A χ^2^ test was employed to test the between-subtype difference of categorical variables. In addition, ANCOVA with age, gender and years of education included as covariates was used to compare continuous variables. For the longitudinal clinical measures (PANSS scales and GAF), a linear mixed model controlling both fixed effects, including subtypes, follow-up time points, age, gender and years of education, and a random intercept (subjects) effect, was built to detect the longitudinal difference (up to 24 months) in psychopathology and global functioning between different subtypes.

### 2.3. Biological function exploration of IT_p_roxygenes f orSCZ

#### 2.3.1. Isolation and culture of rat cortical neurons

To further investigate the function of these IT_proxy genes, especially their effect on neuronal growth, we conducted in vitro studies of these genes on neurons. The cortical neurons were isolated from embryonic day 17-18 Sprague-Dawley rat (purchased from laboratory animal feeding house of Kunming Medical University) brains and cultured as previously described [20,21]. Briefly, the neurons were cultured in Neurobasal® Medium (Gibco, 21103049) (added with 1 × GlutaMAX™ Supplement (Gibco, 35050061), 1 × B-27 (Gibco, 17504044) and 1% heat-inactivated fetal bovine serum (FBS) (Gibco, 10091148)) at first culture day, and the FBS were withdrawn during the following culture days. The half-culture medium was changed every 3 to 4 days. Before neurons culture, a 6 cm cell culture dish was pre-coated with 3 mL 50 μg/mL Poly-D-lysine (PDL) (Sigma, P6407-5MG) for overnight at 37 ^°^C and washed six times using pure water. All neurons without mycoplasma were cultured at 37 ^°^C with 95% air and 5% CO2.

#### 2.3.2. Morphological analysis of dendritic spines

To evaluate the effect of gene over-expression of *ZMAT2* and *PCDHA7* on dendritic spine (a vital component of synapse) formation, the morphological analysis of dendritic spines was carried out as previously described [21, 22]. Briefly, the coding sequence of rat Zmat2 (NCBI Reference Sequence: NM_001135582.1) and Pcdha7 (GenBank: AY573981.1) gene added with HA tag sequence before termination codon were inserted into pCAG-sequencing vectors (EcoRI and NotI) to construct over--expression vectors (pCAG-rat-Zmat2-HA, pCAG-rat-Pcdha7-HA). These vectors were purchased from TSINGKE Biological Technology, and the control and experimental vectors were transformed into DH5α, extracted from a single colony, and verified by Sanger sequencing. The isolated rat cortical neurons were cultured for 14 days in vitro, 3.0 μg experimental vectors and 1.5 μg Venus (expressing GFP proteins are used for labelling spines) were co-transfected into neurons cultured in 6 cm cell culture dishes using 7 μL Lipofectamine 3000 reagent(Invitrogen, L3000015) 72 hours post-transfection. Immunofluorescences were used for staining the spines of neurons which successfully transfected with target plasmids. The primary antibodies are mouse anti-HA tag antibody (1:150, ABclonal, AE008) and chicken anti-GFP antibody (1:1000, Abcam, ab13970). The secondary antibodies are goat anti--mouse 555 (1:500, Invitrogen, A32727) and donkey anti-chicken Cy™2 (1:200, Jackson Immuno Research, 703-225-155). A laser scanning confocal microscope (ZEISS, 880, under 100 × oil lens) was used to scan different layers of spines in neural dendrites which co-expressed GFP and Zmat2-HA or Pcdha7-HA fusion protein. Two secondary or tertiary dendrites of each neuron with a length of at least 100 μm were selected to classify spines (thin, stubby, mushroom, others) and quantify numbers, with each group at least including 30 independent neurons. The images of .czi file with 8 bit format were changed into .tiff by Image J (https://imagej.net/software/imagej/) [22]. The images .tiff file were imported to NeuronStudio (https://m.vk.com/neuron_studio) and combined with the different layer pictures of neural dendrites in Image J for classifying and quantifying spines as previously described [20, 21, 23]. The significant difference between the experimental and control groups was analyzed by a two-tailed Student’s t-test, with the significance threshold value set as 0.05.

## 3. Results

### 3.1. Ten genes were identified as IT-proxy SCZ genes

The expression of 104 genes was found to be associated with IT at a genome-wide significance level (4.85 × 10^−6^, Figure 1). Among them, we found 25 genes to be significantly associated with schizophrenia using MetaXcan; the subsequent analysis using FUSION TWAS (Supplementary material) of these 25 genes identified ten genes to be significantly replicated. The enrichment analysis generated a P value of 5.05× 10^−15^, demonstrating that the findings were not expected by chance. Fig 2 and Table 1 display the association details of 10 IT_proxy genes for SCZ and IT. Consistent with many previous studies [24, 25], Z scores of these ten genes for IT and SCZ, except for (CALN1), are in opposite directions, implying an opposite pleiotropic effect of the same gene on both traits. Using the expression data from BrainSpan [26], we explored the spatial and temporal expression patterns of these IT_proxy genes. As illustrated in Figure 3, the genes were highly expressed in the dorsolateral prefrontal cortex (DLPFC) across nine developmental stages from early fetal to middle adulthood, indicative of the crucial role these genes might play in neuroplasticity and the maintenance of cognitive functions throughout the lifetime. Besides, the genes were also shown to be highly and extensively expressed in other brain areas in the vicinity of DLPFC, such as the orbital frontal cortex and ventrolateral prefrontal cortex, hinting at the vital role of these genes in various cognitive functions.

**Table 1:** Transcriptomic association results of schizophrenia (SCZ) and intelligence (IT) for the IT_proxy genes for SCZ

| Chromosome | Gene name | Ensemble ID | Z score |  | P value |  |
| --- | --- | --- | --- | --- | --- | --- |
|  |  |  | Schizophrenia | Intelligence | Schizophrenia | Intelligence |
| 3 | *PCCB* | ENSG00000114054 | -6.5691 | 4.8826 | 5.06E-11 | 1.05E-06 |
| 3 | *NEK4* | ENSG00000114904 | 5.6499 | -6.5538 | 1.61E-08 | 5.61E-11 |
| 22 | *WBP2NL* | ENSG00000183066 | 5.3859 | -5.9589 | 7.21E-08 | 2.54E-09 |
| 7 | *CALN1* | ENSG00000183166 | 5.0961 | 7.0692 | 3.47E-07 | 1.56E-12 |
| 5 | *PCDHA2* | ENSG00000204969 | 4.999 | -5.5935 | 5.76E-07 | 2.23E-08 |
| 5 | *ZMAT2* | ENSG00000146007 | 4.6295 | -5.8142 | 3.67E-06 | 6.09E-09 |
| 5 | *NDUFA2* | ENSG00000131495 | -4.5648 | 5.8673 | 5.00E-06 | 4.43E-09 |
| 3 | *GLT8D1* | ENSG00000016864 | -4.5578 | 6.4193 | 5.17E-06 | 1.37E-10 |
| 5 | *PCDHA7* | ENSG00000204963 | 4.547 | -5.2023 | 5.44E-06 | 1.97E-07 |
| 2 | *ALMS1-IT1* | ENSG00000230002 | 4.195 | -5.5145 | 2.73E-05 | 3.50E-08 |

**Fig. 2:**
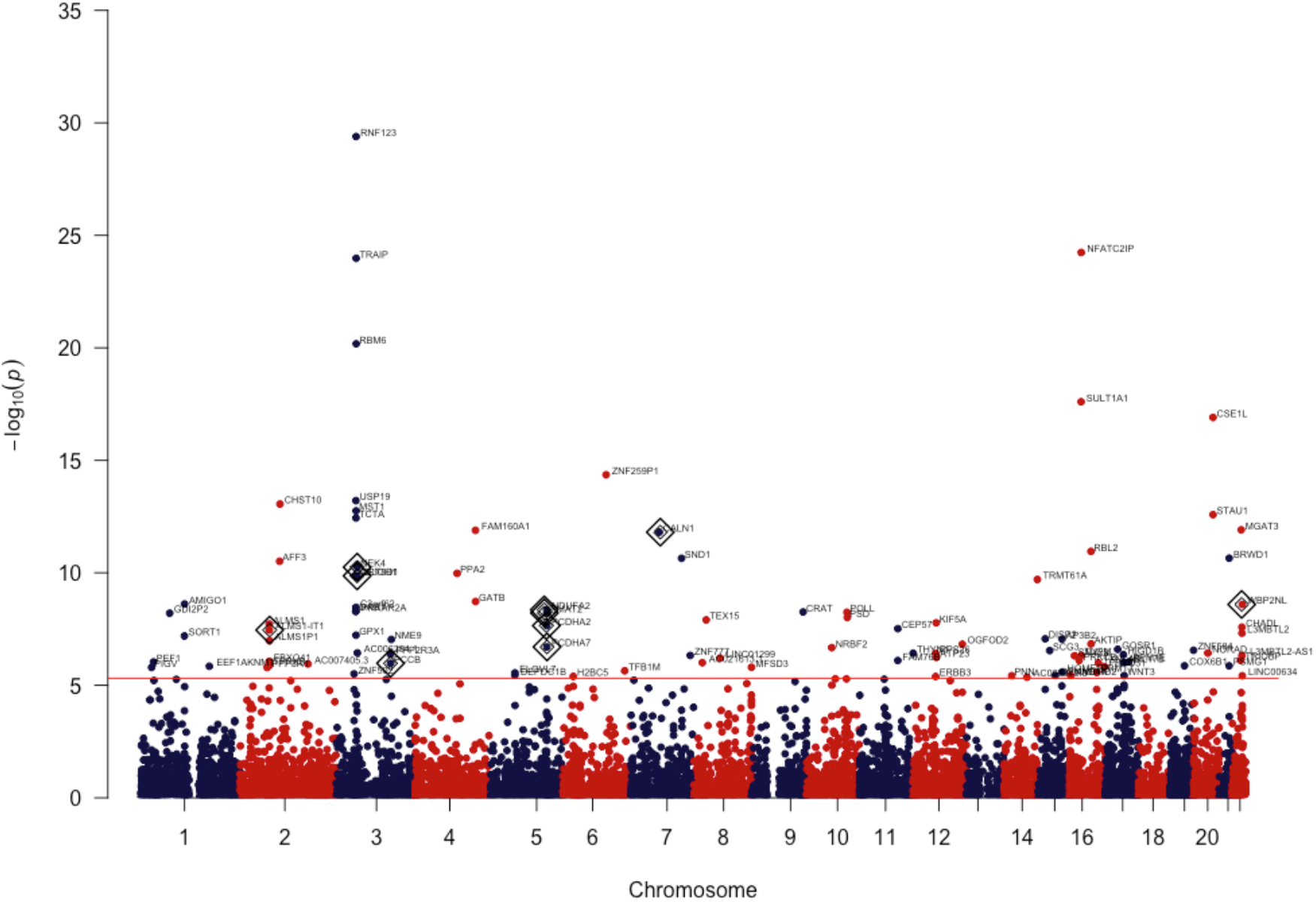
Manhattan plot of association results of intelligence in DLPFC, annotated dot being one the association of which reached a genome-wide significance level and the highlighted one with concentric diamond being ones significantly associated with schizophrenia using PPM

**Fig. 3:**
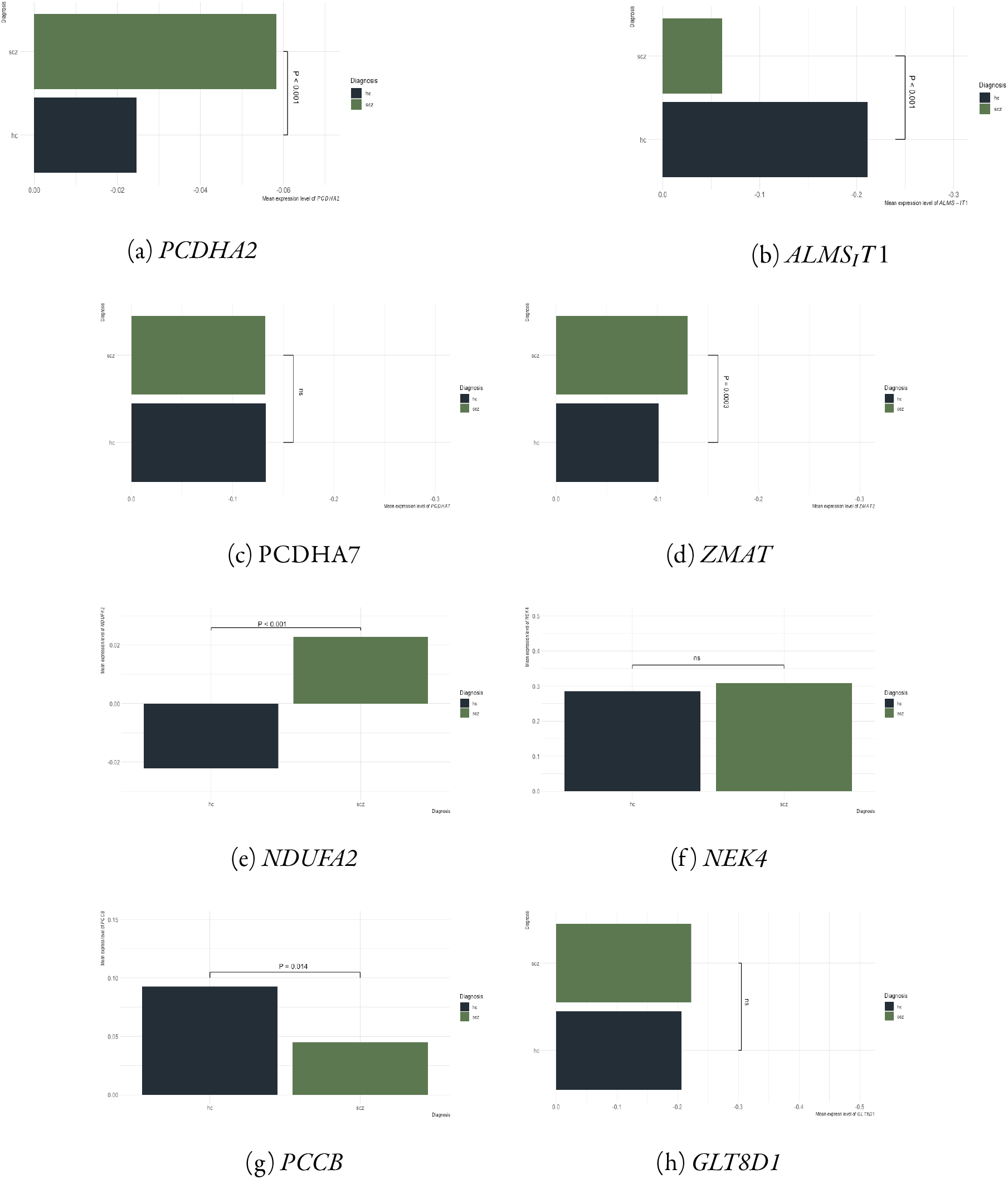
GReX comparison of ten IT_proxy genes for SCZ between first-episode, drug-naive patients with SCZ and healthy controls

### 3.2. Real-world data analysis: four biotypes emerging from expression matrix of 10 IT_proxy genes

Following the GReX prediction of these ten genes, we compare the GReX level for each gene between patients and healthy controls. The results are displayed in Fig 3. Among these ten genes, *GLT8D1, NEK4*, and *PCDHA7* did not differ significantly between patients and controls, and the comparison of another two genes, *WBP2NL* and *CALN1*, were invalidated by the fact that their expression could not be derived from the control group.

The clustering analysis using the t-distributed stochastic neighbour embedding (TSNE)revealed four distinctive clusters (biotypes) existing in the patients when the hyper-parameters, perplexity and iteration steps were set as 25 and 5000, respectively Ω Fig 4ΩL.

**Fig. 4:**
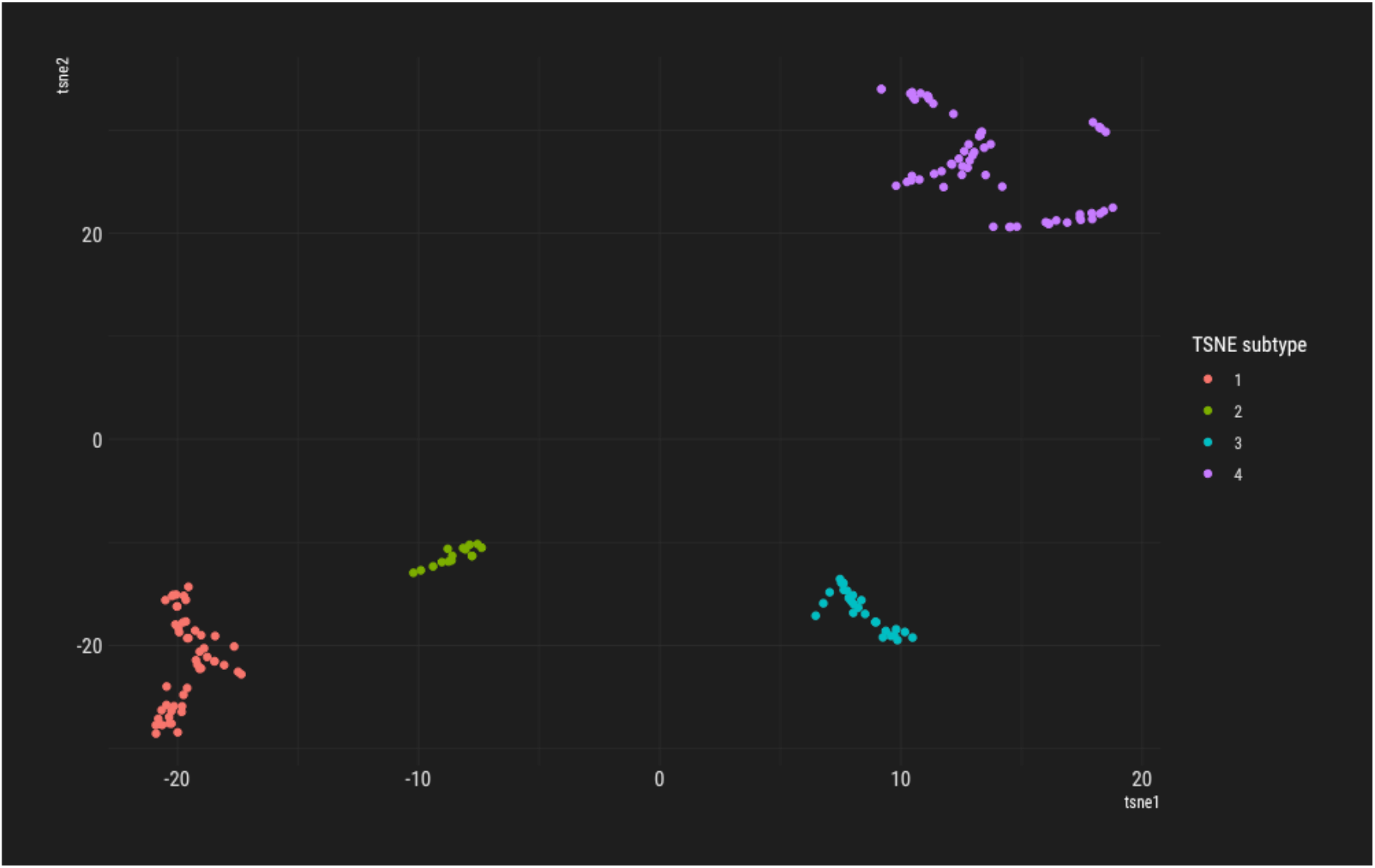
Cluster delineated in the patients group using TSNE (perplexity and iteration steps set as 25 and 5000, respectively)

### 3.3. Sensitivity analysis

Given that three genes did not differ significantly in their expression between cases and controls and the expression of another two genes could not be derived in the control group, we reran the clustering analysis by excluding these five genes. The result supported our primary findings of 4 clusters, with most individuals staying in the same cluster as one generated using the expression matrix of 10 genes. Besides, the cluster analysis using another clustering algorithm, KMEANS, replicated our primary cluster numbers with 100% consensus.

### 3.4. Profiling of each biotypes revealed a notable heterogeneity in SCZ with regard to cognition, neuroimaging, psychopathology and longitudinal disease course

No significant difference was detected in demographic features, including age, gender and years of education, between biotypes. However, the four biotypes differed significantly in working memory capacity, tested using the module in CANTAB, delayed matched to sample (DMS). As displayed in Fig 5, biotype two performed more poorly than the other three biotypes, with more errors (DMS_PEGC: DMS Prob error given correct), lower correct rate (DMS_TC: DMS Total correct; DMS_TC_A: DMS Total correct (all delays); DMS_TC0D: DMS Total correct (0 ms delay); DMS_TC12D: DMS Total correct (12 ms delay)).

**Fig. 5:**
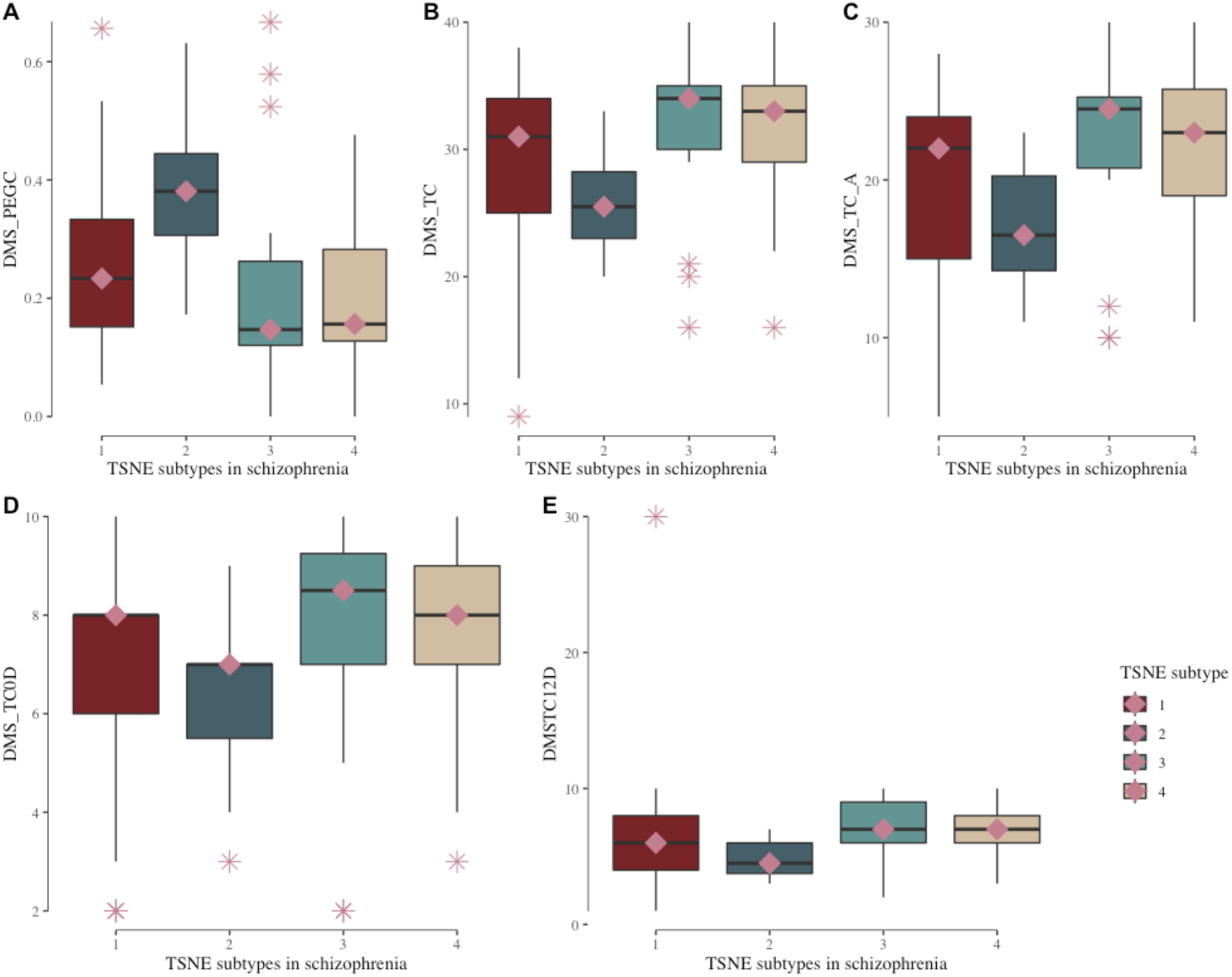
Working memory capacity comparison between different biotypes and controls using delayed matched to sample (DMS)

The results of GMV comparisons between four biotypes are demonstrated in Fig 6. Five brain regions showed significant differences in the GMV between biotypes and survived multiple comparisons using FDR (*P*_*lLimbicA*_*TempPole*_2_*FDR*_ = 0.002967, *P*_*rLimbicB*_*OFC*_1_*FDR*_ = 0.0357, *P*_*rLimbicA*_*TempPole*_2_*FDR*_ = 0.0357, *P*_*rContA*_*IPS*_1_*FDR*_ = 0.0357, and *P*_*rContB*_*PFClv*_1_*FDR*_ = 0.0181). The differentiation patterns of four biotypes and healthy controls in these five brain regions implied the complexity and heterogeneity at the neural structural level in schizophrenia:

**Fig. 6:**
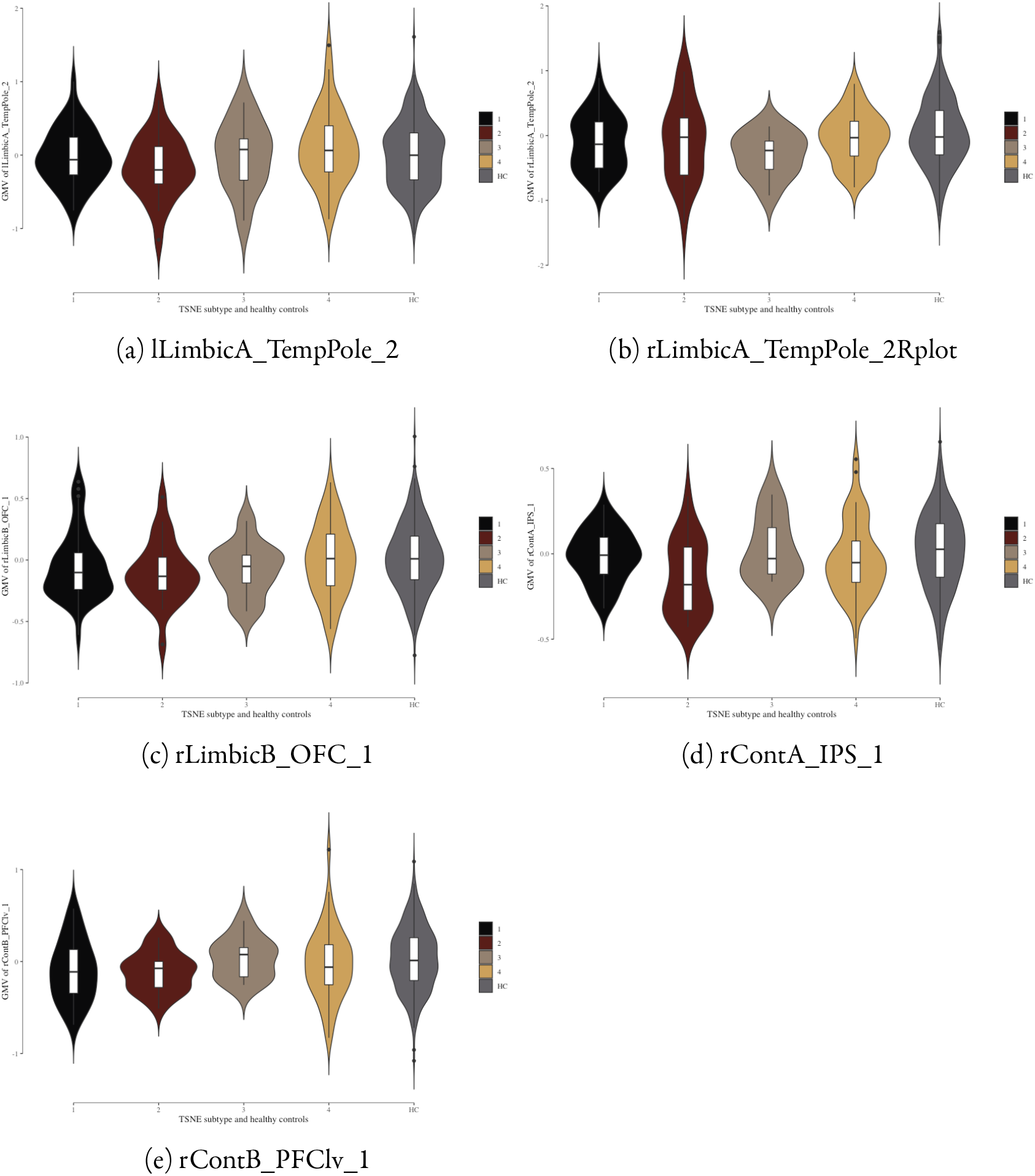
Gray matter volume (GMV) comparison between different biotypes and healthy controls

1. Compared with healthy controls, biotype two showed significantly less GMV in all identified brain regions, the one with the least GMV out of 5 groups in lLimbicA_TempPole_2, rLimbicA_TempPole_2, rLimbicB_OFC_1, rContA_IPS_1 and rContB_PFClv_1.
2. The GMV of healthy controls, intriguingly, was not consistently the largest among all five groups, including four disease biotypes and healthy controls except for rLimbicA_*T*_ *empPole*_2_.
3. The GMV of healthy controls in the remaining four brain regions stood somewhat between that of the first two biotypes and that of the last two biotypes.

Besides comparing the GMV of individual brain regions, we also investigated the pattern difference between different groups in the correlation network of the five brain regions identified above. As demonstrated in Fig 7, the correlation network of biotype two, in contrast with other subtypes and healthy controls, showed a notably weaker correlation between lLimbicA_TempPole_2 and rLimbicA_TempPole_2. In addition, the correlation of rContB_PFClv_1 with other brain regions was more negative in biotype two than in other biotypes and healthy controls, hinting at a more pervasive and persistent desynchronization and decoupling among the cognition-associated brain regions in a subgroup of patients with schizophrenia.

**Fig. 7:**
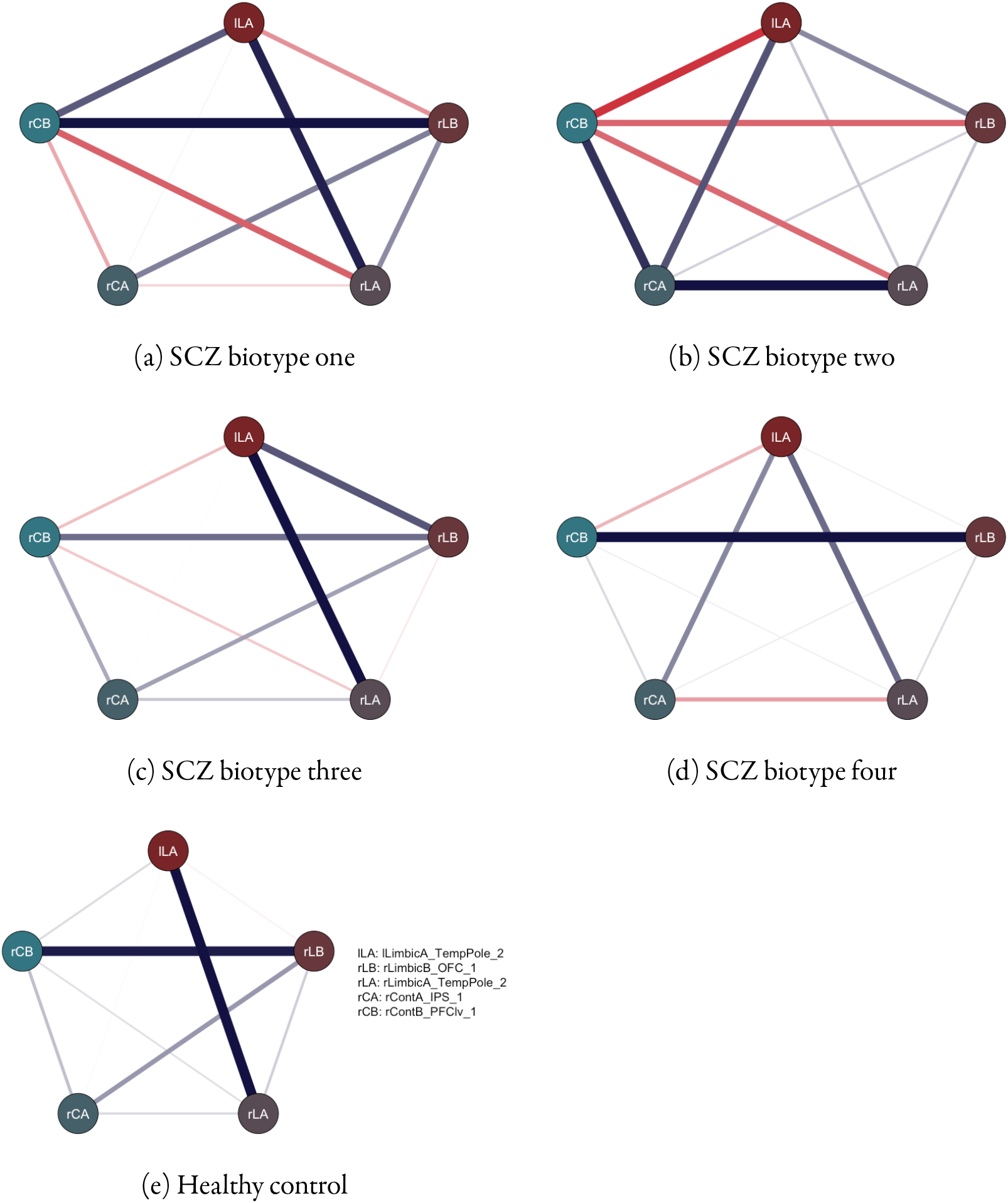
Comparison in structural network of five identified brain regions between different biotypes and healthy control

The longitudinal analysis of the PNASS scale and GAF indicated that while no difference was detected between biotypes in positive, negative and general composite scores, biotype two scored significantly higher than the other three subtypes in the PANSS item N4 (“Passive/apathetic social withdrawal”, P = 0.023).

### 3.5. IT_proxy genes implicated in neuronal plasticity

The dendritic spine is a crucial component of the synapse, which plays essential roles in schizophrenia and intelligence, and previous studies have repeatedly reported significant effects of schizophrenia risk genes on dendritic spine morphogenesis, such as *NEK4* and *GLT8D1* in the present gene list [27, 28]. Hereby we used rat cortical neurons as the cell model to conduct morphological analysis to evaluate the additional risk genes confer on the dendritic spine density. We selected one risk gene (*PCDHA7*) from the protocadherin alpha gene cluster, which has shown essential roles in neuronal development and functions [29,30]; and another risk gene (*ZMAT2*), the function of which in neuron is still unclear but exhibits higher mRNA expression in the brains of schizophrenia patients compared with normal controls. Our results showed that over-expressing *Zmat2* gene significantly increased the density and ratio of stubby spines compared with that in control groups (Fig 8: A-C). In contrast, the density and ratio of mushroom spines were significantly decreased in rat Pcdha7 overexpressing neurons (Fig 8: D-E). These results indicated that *ZMAT2* and *PCDHA7* might partcipate in dendritic spine morphogenesis relevant to pathogenesis of schizophrenia, despite the precise mechanisms may be distinct.

**Fig. 8:**
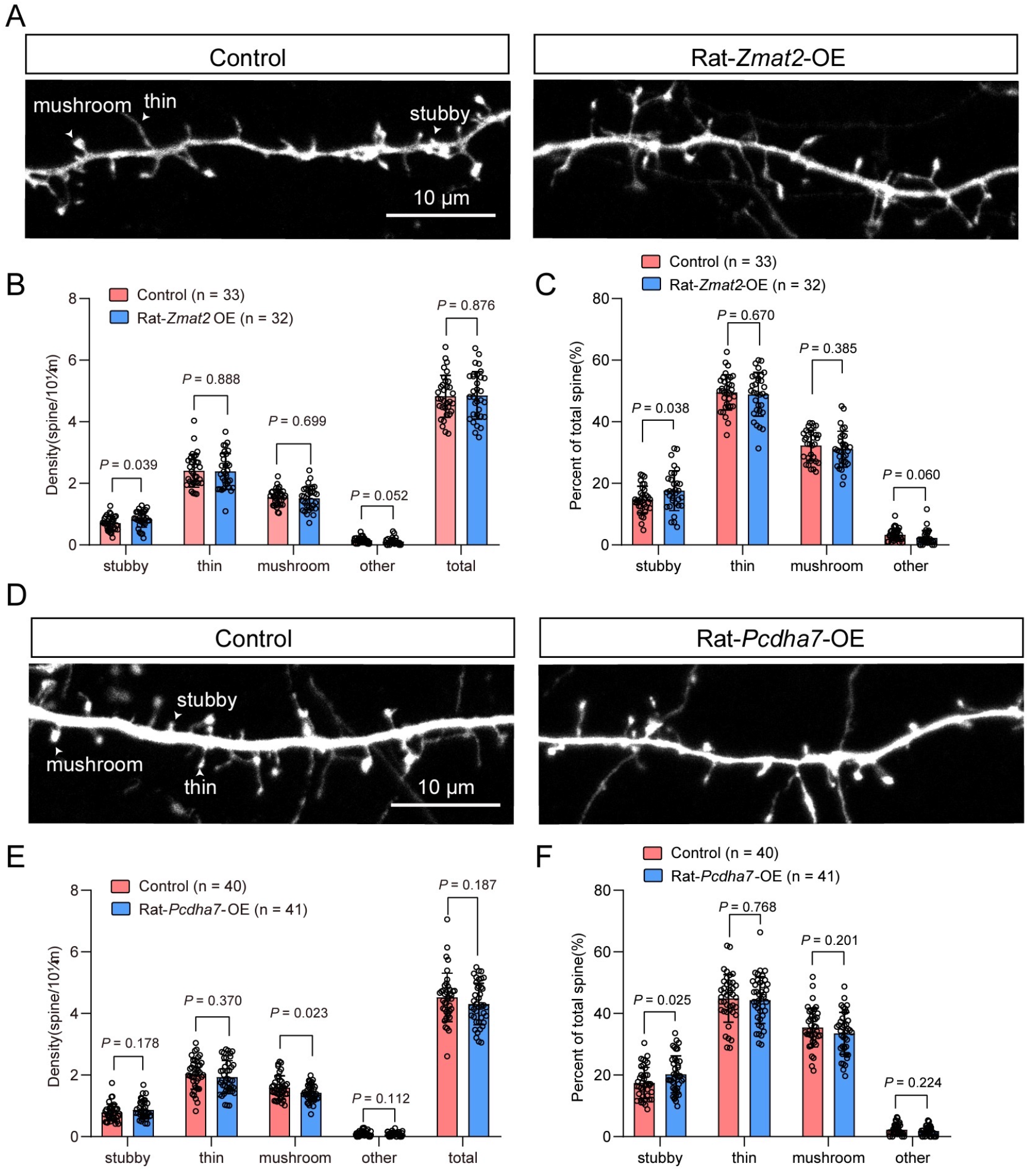
Comparison of dendritic spines between control group and over-expressing *ZMAT2*(A-C) and *PCDHA7* (D-E), respectively

## 4. Discussion

Our present study used intelligence (IT) as a proxy phenotype to detect genes showing genetic dependence between IT and schizophrenia(SCZ). To identify genes in a more spatially fine-grained way, we chose gene expression value in DLPFC as our instrument. We found ten genes, the expression of which in DLPFC displayed a genetic dependence between IT and SCZ (IT_proxy genes for SCZ). Further exploration in an independent sample indicated an efficacy of ten genes herein to cluster SCZ patients into four biotypes, with each of these biotypes demonstrating pathological changes in cognition, brain structures and psychopathology.

Among ten IT_proxy genes for SCZ, *ZMAT2* (Zinc finger matrin-type protein 2) is a 5-member family in humans, in which all the encoded proteins contain zinc finger domains. *ZMAT2* is highly conserved in humans and non-human primates [31]. An inquiry in the GTEX portal showed that *ZMAT2* is highly expressed in various brain regions, yet there have not been many studies on its biological function in the brain. Our study found ZMAT2 to significantly increase the density and ratio of dendritic stubby spines. Dendritic spines are tiny protrusions from dendrites and form functional contacts with other neurons; they are highly plastic and can adjust their shape and size in response to neuronal activities [32]. Many studies have shown that the dendritic spines are the sites for memory formation and storage. Dendritic spines can manifest in many shapes and sizes, with mushroom, thin, stubby and filopodia being their major shape categories. Stubby spines tend to predominate the post-synaptic component in the postnatal age and gradually disappear into adulthood [33]. Helm et al., in their study, annotated more than 47,000 spines for more than 100 synaptic targets and observed that stubby spines, long recognized as the immature spines, owned similarity with mushroom spines, the mature ones, in both protein numbers and topologies, but not in the correlation between proteins and synaptic strength [34]. Our study suggests that *ZMAT2* could increase the plasticity of stubby spines, possibly in the early stage of life.Considering the neurodevelopmental nature of SCZ, our study provides another strategic angle to investigate the mechanism underlying the cognitive deficits of SCZ. Intriguingly, in contrast with the role of *ZMAT2* in the maintenance of stubby spines, our study found another IT_proxy gene for SCZ, *PCDHA7*, to be able to decrease the density and ratio of mushroom spines. *PCDHA7* (Protocadherin alpha 7) is a calcium-dependent cell-adhesion protein involved in cell adhesion and recognization. PCDHs, including PCDHA2 found in the current study, belong to the cadherin superfamily and are implicated in regulating cellular contacts. PCDHA7 is one member of clustered subfamily highly expressed in multiple brain regions, including the cortex, hippocampus, cerebellum, etc. PCDHs are calcium-dependent and were found to be critical in both synaptic pruning and stabilization. Our study, arguably for the first time, deciphers the function of PCDHA7 at the cellular level. Since mushroom spines are the primary dendritic spines in adulthood, their maintenance plays an essential role in learning and other cognitive functions. Its disruption could stall the normal neurodevelopmental process leading to psychiatric illnesses such as SCZ [35].

Besides, *ALMS1-IT1* was found by our study to show a notable genetic dependence between IT and SCZ and a significant difference in predicted expression between cases and controls in an independent sample. *ALMS1-IT1* is an RNA gene and widely expressed in brain tissues. One study linked the expression of *ALMS1-IT1* to NF-kB signalling in neuroinflammation in the wake of ischemic cerebral injury. Cui-Ping et al. used an integrative analysis to pinpoint causal genes for SCZ, and they identified *ALMS1* as one of the causal genes for SCZ, which was validated using eQTL and SMR [36]. These results further validated our findings that ALMS1 might confer risk of SCZ by disrupting the neuroinflammatory pathway involved in cognition [37]. Additionally, *NDUFA2* and *PCCB* encode proteins which are vital components in the energy process in mitochondrial. Accumulating evidence recognizes mitochondrial dysfunction as a major source of the pathogenesis of SCZ [38, 39].

Transcriptomic enrichment analysis of differently expressed genes (DEGs) in the upper-layer GABAergic interneurons at the DLPFC of the postmortem brain of schizophrenic patients uncovered the down-regulation in energy metabolism. It could be speculated that the effect of NDUFA2 and PCCB-involved pathways on energy processes could be in opposite directions in IT and SCZ and that restoring such a pathway could be a novel treatment target for SCZ.

Our current study detected a specific biotype in schizophrenia which linked working memory deficit and reduced GMV to the core psychopathologic trait, especially that of negative symptoms. Many studies have acknowledged that the negative symptoms could be underpinned by a patho-etiology discrete from that underpinning other symptom syndromes of SCZ [40]. While the differentiation pattern falls short of discretion between each group, including healthy controls in the GMV of individual brain regions, the structural correlation network between chosen brain regions illustrates a clearer pattern between different groups. In particular, the correlation between lLimbicA_TempPole_2 and rLimbicA_TempPole_2 is the strongest in healthy controls and the weakest in biotype2. Many studies have shown that the interaction between various brain regions and thousands of neurons subserves most of our living experiences. Temporal poles are of intrinsic interest in schizophrenia and are critical in episodic and semantic memory [41, 42]. The biotypes detected in our study showed various degrees of working memory deficits, and we argue that such deficits might arise from structural connectivity dysfunction with the temporal poles as its hub.

## 5. Limitation

Although our results arise from rigorous study planning and implementation, we are fully aware of a few limitations in our study. First, the reference data used in the prediction of genetic expression in our sample of Chinese Han ancestry comes from populations of European descent. Whereas population structure remains a major confounding factor in genetic association analysis, some studies also prove that the information gleaned from one population could be employed in another population [43]. While our results shed new light on the cognitive deficits of schizophrenia, a transcriptomic database involving a large sample of the Eastern Asian population should be guaranteed in the future to increase the accuracy of prediction. Further, there is a notable dropout rate in our follow-up data, only forty-four patients finished the follow-up at 12 months, and twenty patients stayed at 24 months. Attrition remains a main issue in prospective studies. While our study provides a trend worthy of further replication and exploration, a larger longitudinal study in the future is needed to replicate our primary findings.

## 6. Conclusion

For the first time, our study used intelligence (IT) as a proxy phenotype to reduce the heterogeneity of schizophrenia (SCZ). The results showed that ten genes, the expression of which in the dorsal lateral prefrontal cortex (DLPFC), were associated with IT and also played a potentially core role in the pathogenesis of SCZ, especially the cognitive deficits of SCZ. Further, using such knowledge of gene expression in specific brain areas could reduce the heterogeneity of SCZ and help identify subgroups with similar pathophysiological changes and clinical profiles. As shown in our study, four biotypes arose from the expression data of these ten genes in DLPFC, and each of these biotypes exhibits distinctive cognitive capacity, gray matter volume (GMV) and psychopathology. In addition, further exploration at the cellular level implicates these genes in dendritic spine growth, indicative of their role in neuroplasticity.

## Data Availability

All data produced in the present study are available upon reasonable request to the authors

